# Modified Technique for Dissection of Working Space in Retroperitoneal Laparoscopic Surgery

**DOI:** 10.1101/2023.06.18.23291359

**Authors:** Xiaofeng Xu, Wen Cheng, Zhe Liu, Changjie Shi, Jianping Da, Xiumin Zhou, Jingping Ge

## Abstract

**Introduction:** The existing techniques to creat working space for retroperitoneal laparoscopic surgery are often accompanied by air leakage or poor visibility due to blood and fat tissues smudging during dissection.This single-center experience describes a modified dissection technique to create a retroperitoneal working space during laparoscopic surgery.

**Materials and Methods:** From May 2021 to December 2022, we performed a modified dissection technique to create a retroperitoneal working space prior retroperitoneal laparoscopic surgery in 47 patients. During the procedure, laparoscopic dissection is successively performed. The retroperitoneum is initially accessed by puncturing the trocar through a 10-mm transverse skin incision in the midaxillary line. Under endoscopic monitoring, the tip of the trocar is adjusted to a relative avascular layer between the transversus abdominis muscle and the pararenal fat. Laparoscopic dissection is performed to develop until the working space is fully established. All data referring to patient demographics, surgery, dilation-related complication, and perioperative outcomes were collected retrospectively.

**Results:** In all cases, a satisfactory retroperitoneal space was created for surgery. The median time of creating retroperitoneal working space was 6 (IQR:5,7) minutes. No dissection-related complications were noted within a median follow-up period of 9 (IQR:7,15) months.

**Conclusion:** Modified retroperitoneal dissection with laparoscopy is a safe, simple, effective, and minimally invasive technique. It provides an adequate working space and an excellent view without obvious bleeding.

## Introduction

Creating an appropriate working space within the retroperitoneum is an essential prerequisite for retroperitoneal laparoscopic surgery. Early attempts at creating adequate space were mostly unsuccessful until the balloon dilation technique was first described by Gaur in 1992[1]. Consequently, the use of this technique has become increasingly popular worldwide. However, this technique utilizes an open surgical approach and air leakage is an inevitable problem after dilation because the incision for digital dissection is larger than the diameter of the trocar [2].

To overcome the complications associated with the existing techniques, we have implemented a modified dilation technique at our institution. This technique achieves retroperitoneal access by direct trocar puncture through a 10-mm incision. The laparoscopic dissection technique is used to dissect pararenal fat (PF) from the transversus abdominis muscle (TAM). Herein, we report our experience with this modified dilation technique.

## Materials and methods

### Patients

We have successfully applied this modified dilation technique in 47 cases of benign and malignant diseases of the upper retroperitoneum at our institution. Informed consent was obtained from all the patients. Perioperative data were collected retrospectively after obtaining approval from the Institutional Review Board of Jinling Hospital. Follow-up data were obtained by reviewing patient charts or via telephone calls.

### Step-by-step technique

After inducing general anesthesia, the patient is placed in a full-flank position with the kidney bridge elevated. The surgical table is flexed to increase the distance between the ribs and iliac crest. A 10-mm transverse incision is made in the midaxillary line, approximately 2 cm above the iliac crest (Fig. 1a). A 10-mm trocar is punctured perpendicularly through the muscular layers and transverse fascia and a 10-mm, 30-degree laparoscope is inserted into the trocar. The laparoscopic view is turned to 30-degrees up. Under laparoscopic monitoring, the trocar is adjusted until its tip is within the layer between the TAM and the PF (Fig. 1b). This layer consists of semi-transparent loose and flimsy fibroareolar tissue and is relatively avascular. The eyepiece of the laparoscope is subsequently pushed downward. Using this angle ensures that endoscopic vision remains in the proper layer (Fig. 1c). Thereafter, to-and-fro and side-to-side laparoscopic movements are performed to separate the fibrous septae, and the PF is bluntly detached from the undersurface of the flank abdominal wall. Accordingly, a working space is created between the TAM and PF.

**Fig. 1.**
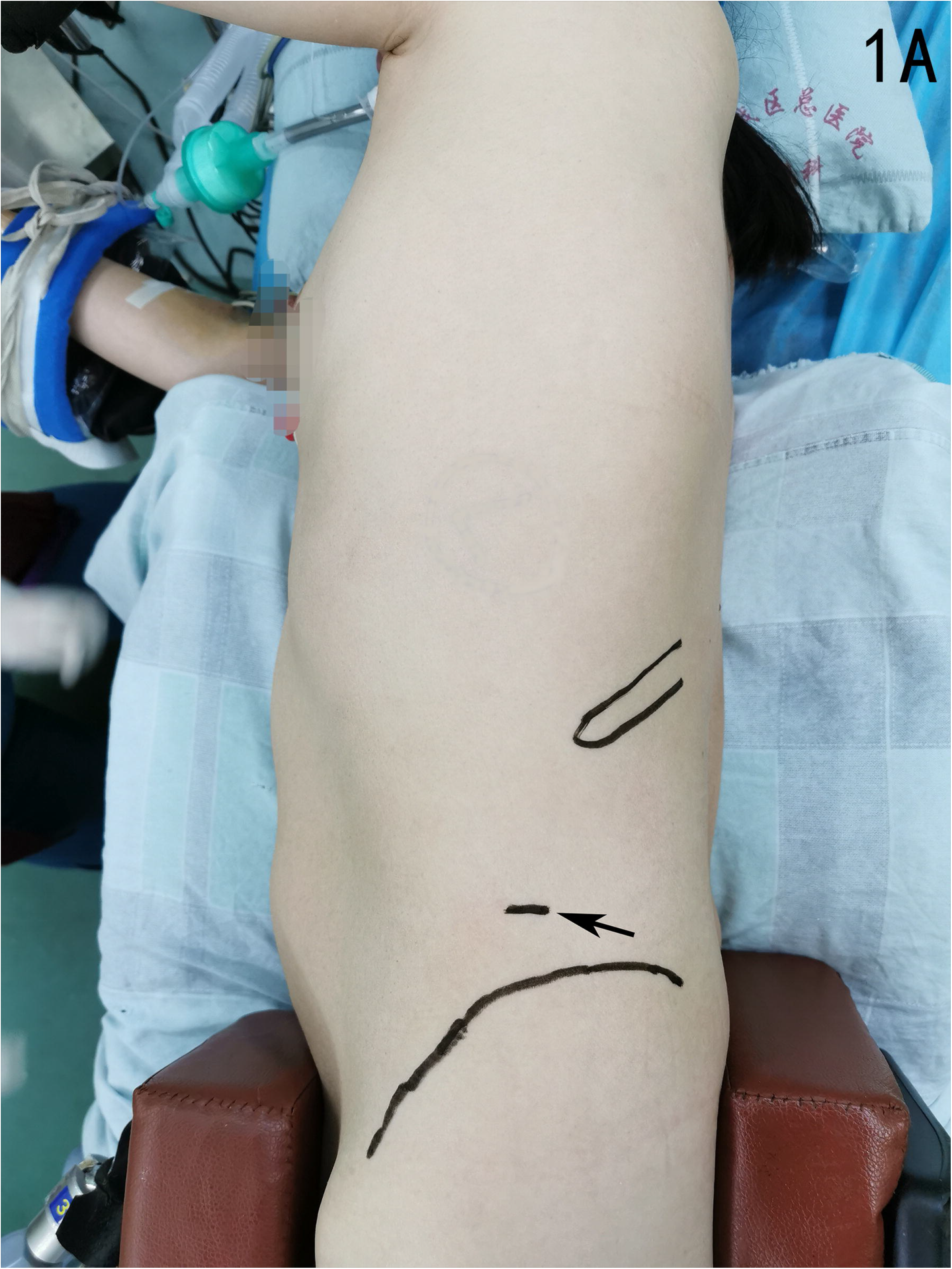
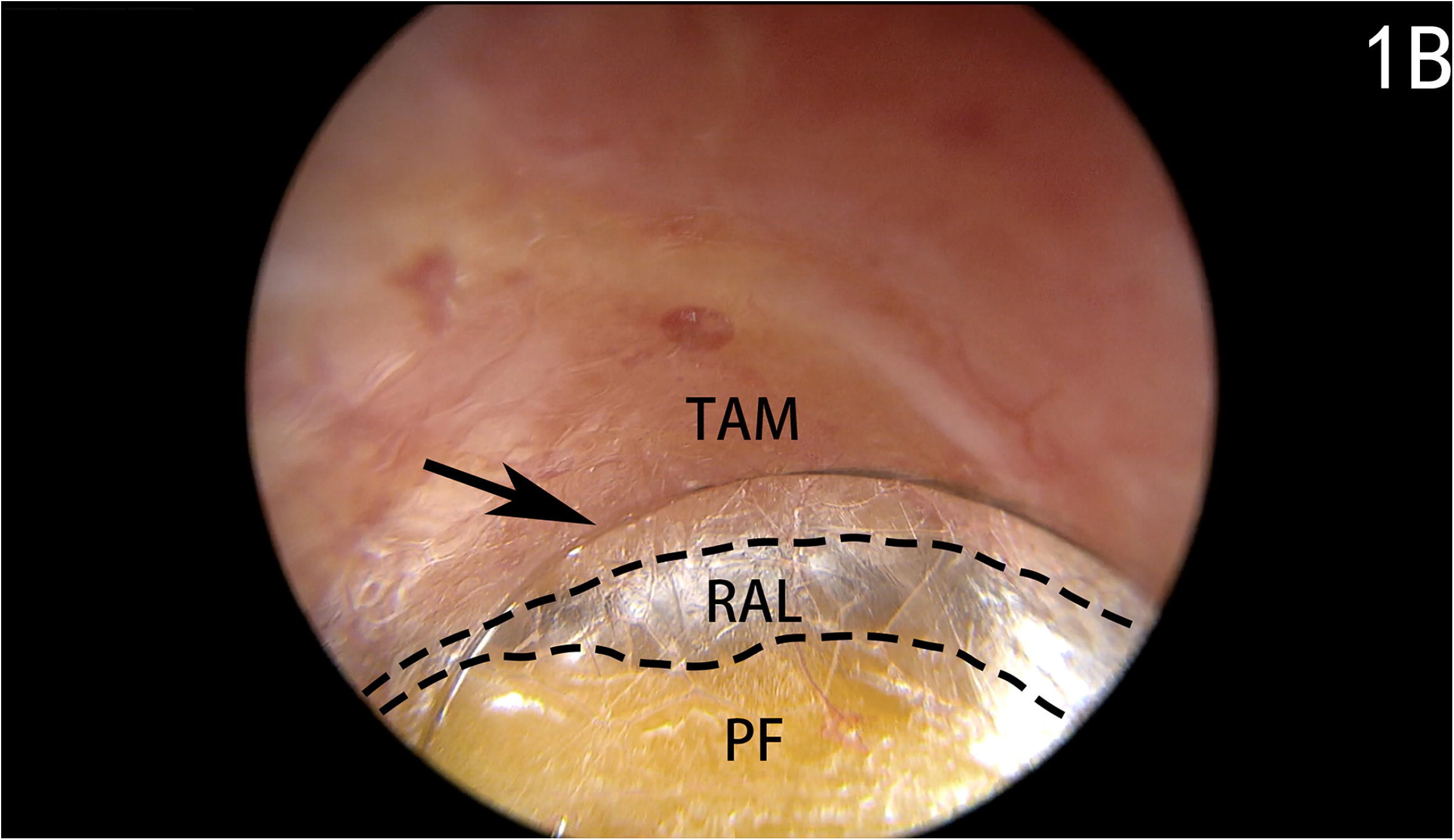
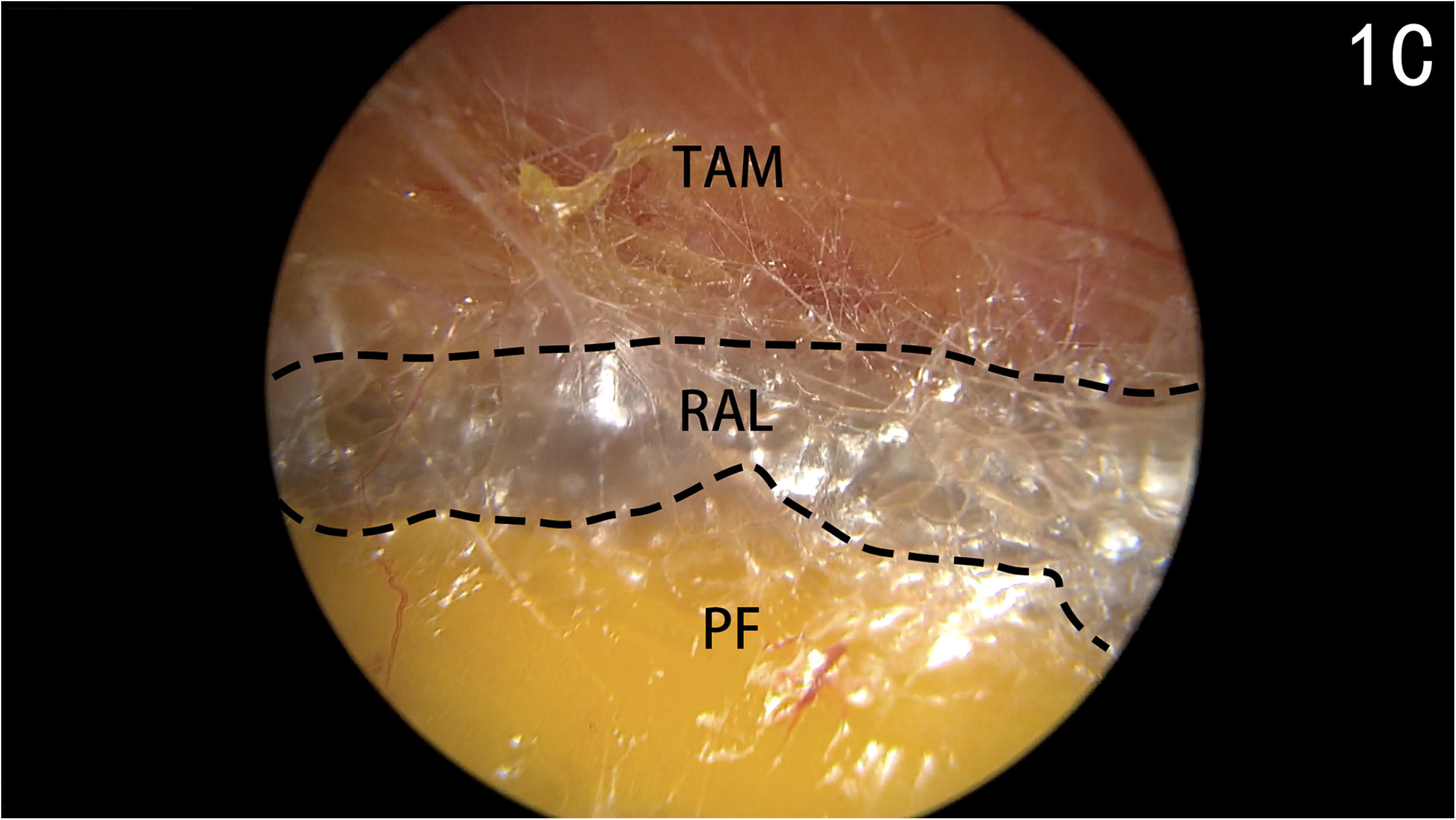
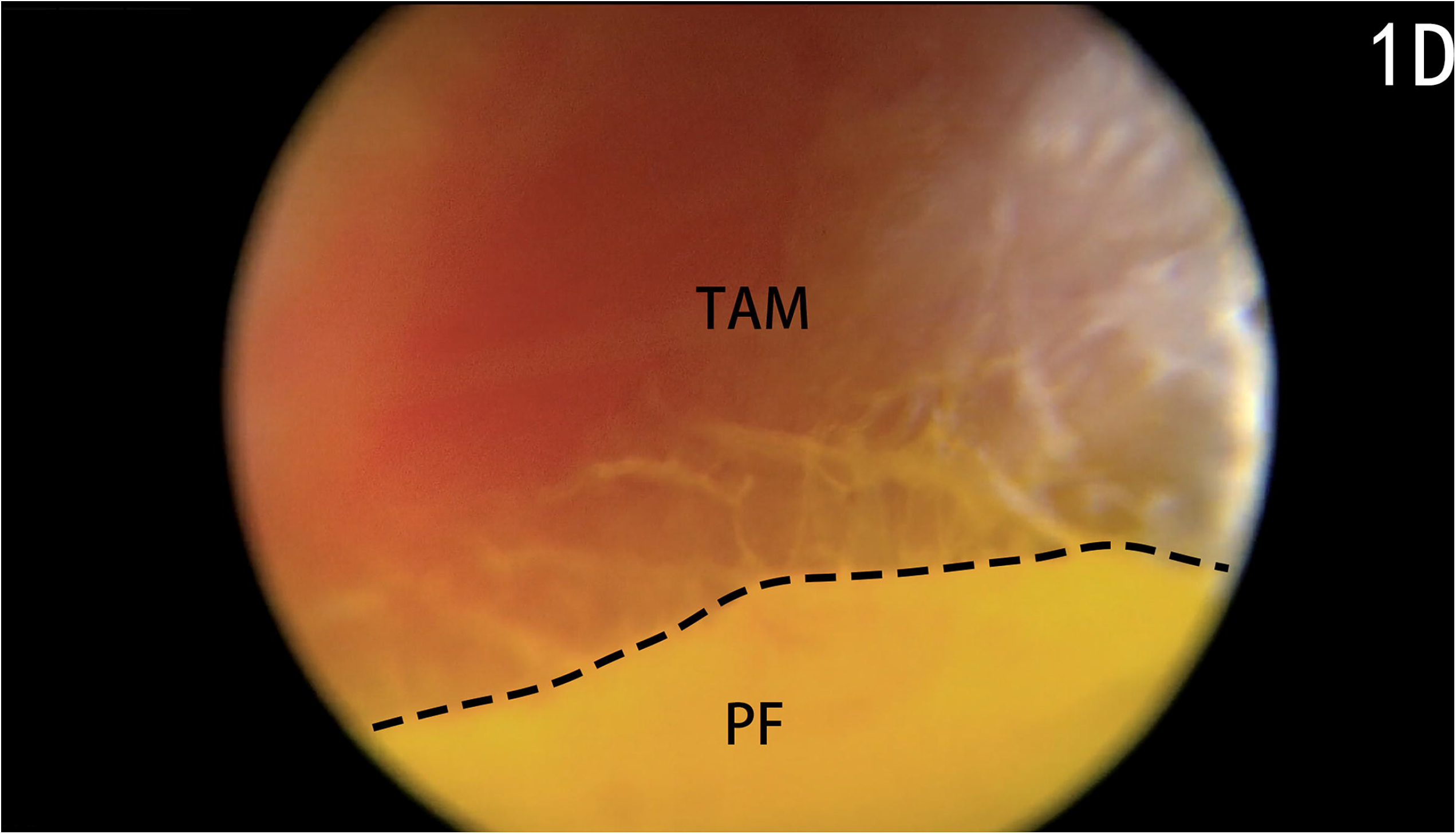
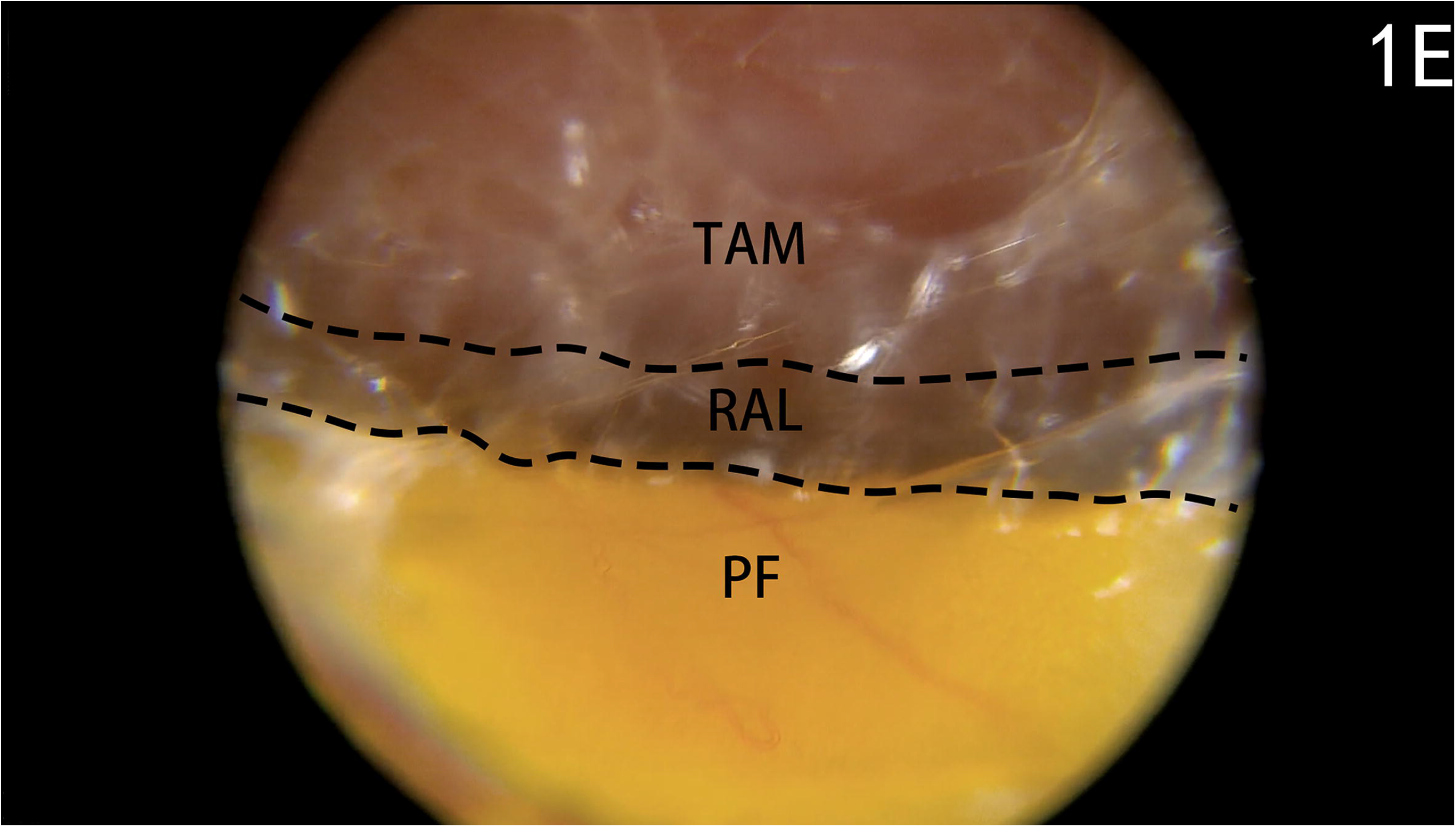
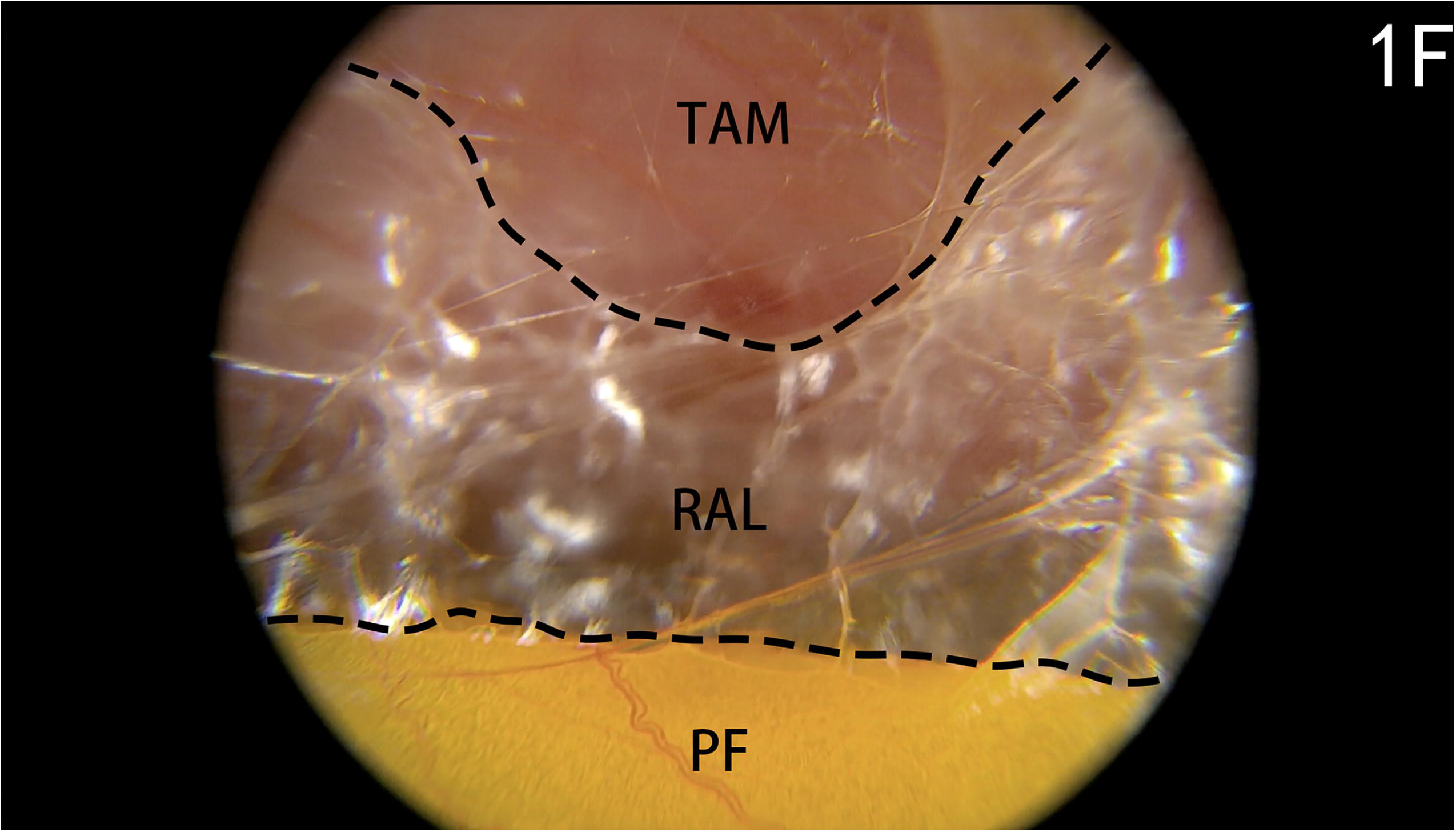
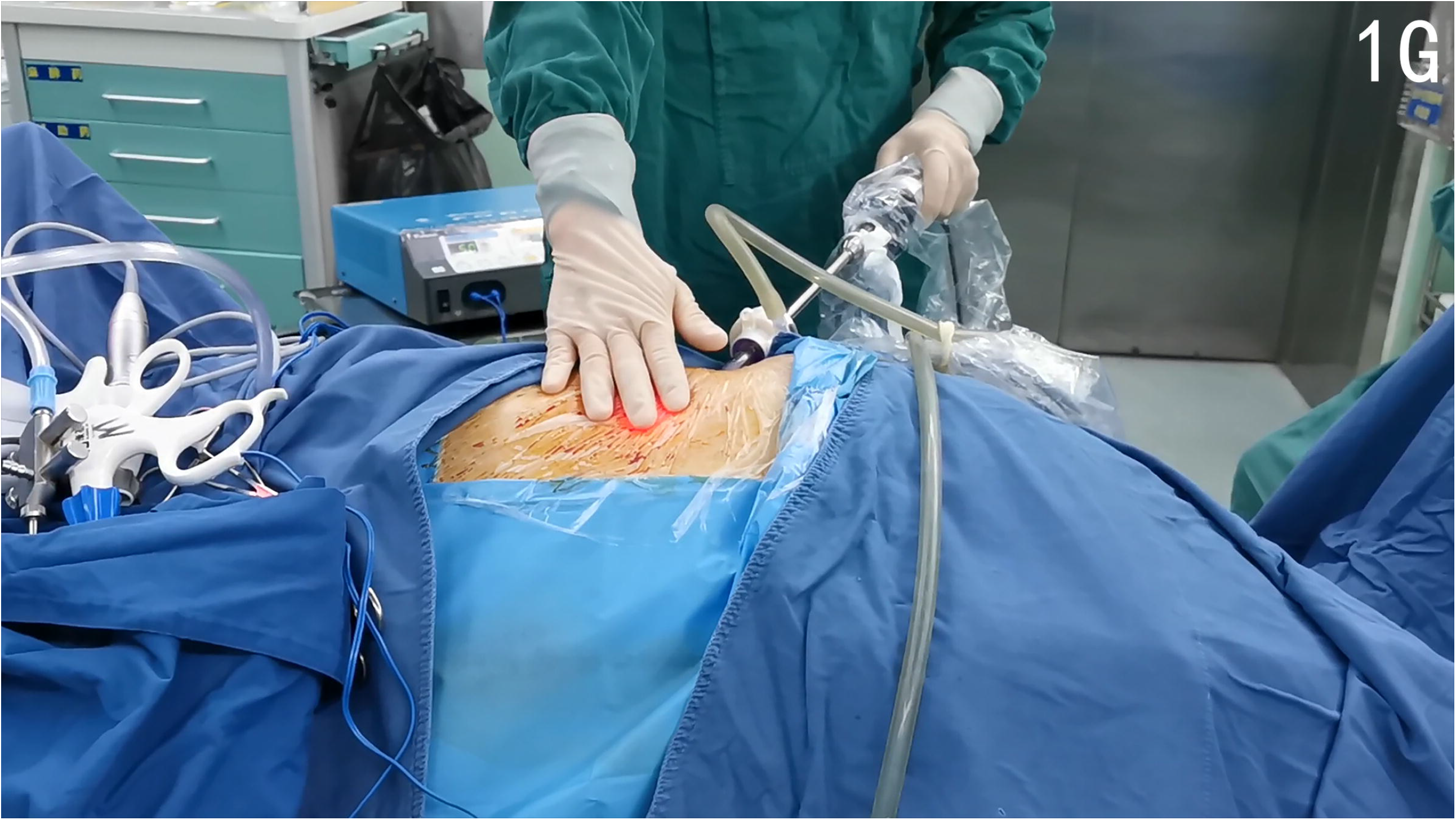
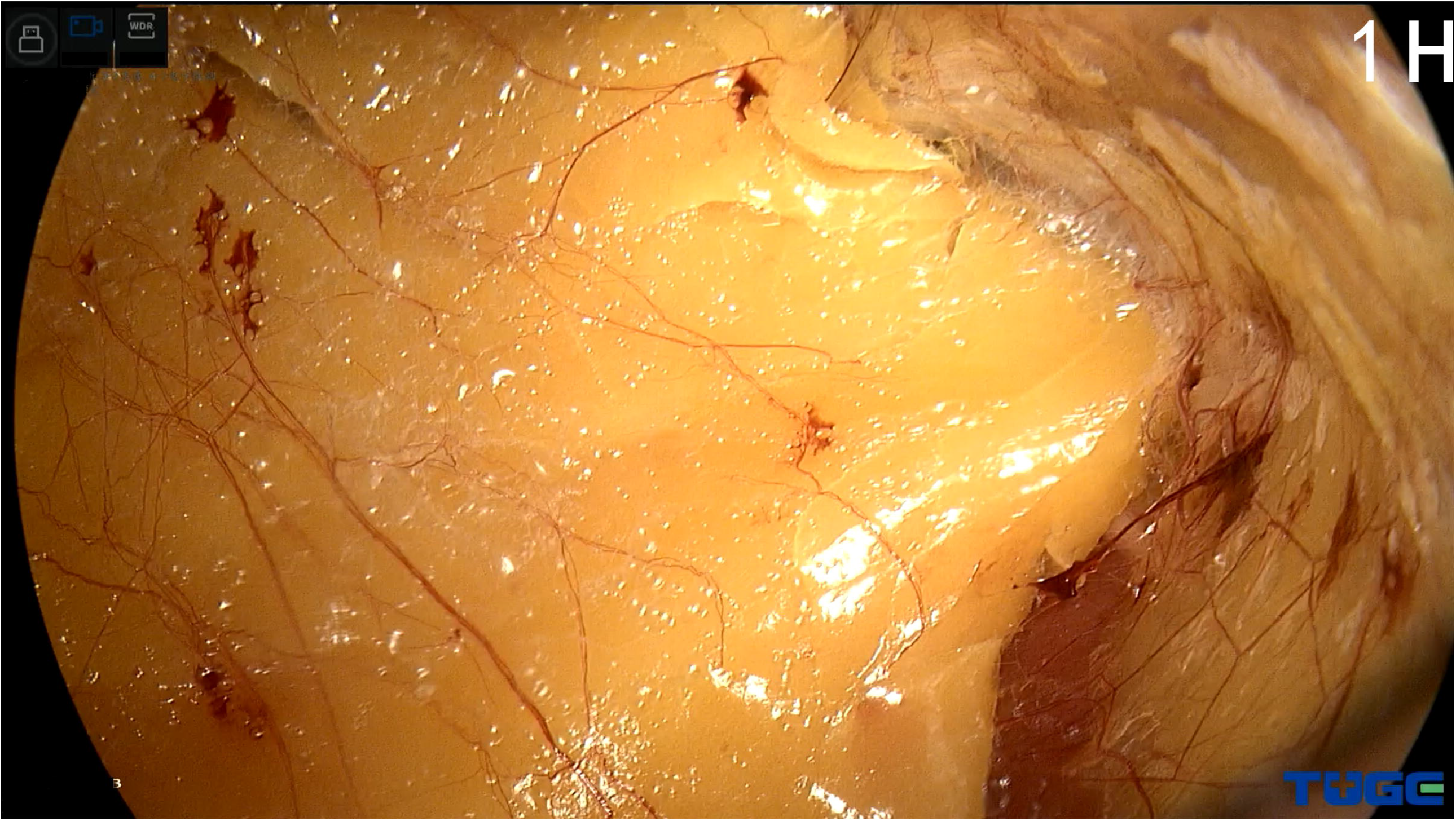
Key procedures of the modified dissection technique for creating an adequate retroperitoneal working space. **a** The patient in a full-flank position. The first trocar is punctured perpendicularly through a 10-mm incision (black arrow) in the midaxillary line, approximately 2 cm above the iliac crest. The 12th rib and iliac crest have been marked. **b** Telescopic view of the 30-degree up from the first trocar. The trocar tip (black arrow) is adjusted to the relatively avascular layer (RAL) between the transversus abdominis muscle (TAM) and pararenal fat (PF). **c** Laparoscopic dissection within the RAL. **d**,**e**,**f** The laparoscopic view is out of focus during dissection. The PF is yellow, the TAM is red, and the RAL is semi-transparent. These characteristics can be regarded as landmarks. **g** Strong light from the laparoscope transilluminates the muscle and forms a light spot on the skin. Movements of the laparoscope are palpable by hand simutaneously. **h** The working space is fully established. PF is mobilized en bloc from the transversalis fascia. A clear laparoscopic view without hemorrhage is observed

**Fig. 2.**
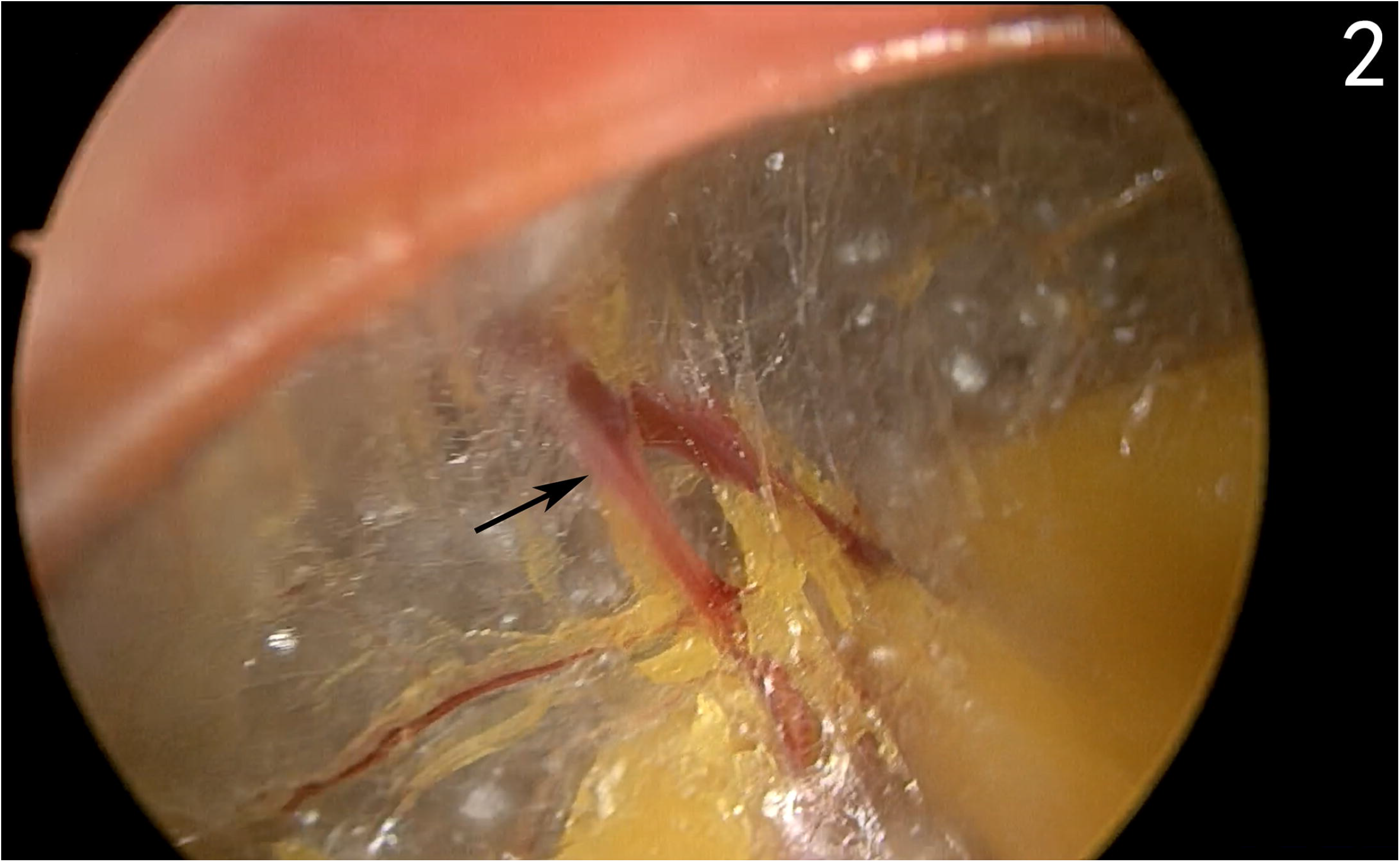
Perforator vessel in fibroareolar tissue (black arrow)

There are several technical tips to ensure that laparoscopic dissection remains in the appropriate layer while using this technique. (1) Laparoscopic focus should be adjusted promptly when object distance is too small which will lead to obscure view. (2) The retroperitoneal fat is yellow, the TAM is red, and the fibroareolar tissue is semi-transparent. As the laparoscopic view is out of focus during dissection, these characteristics can be regarded as landmarks (Fig. 1d-1f). (3) Strong light from the laparoscope can transilluminate muscle and form a light spot on the skin. This can aid in locating the tip of the laparoscope (Fig. 1g). (4) The tip of the laparoscope should be pressed onto the undersurface of the flank abdominal wall and the wall will be tented during laparoscopic dissection. Thus, the movements of the laparoscope under the skin are palpable (Fig. 1g).

The dissection area extends from the anterior axillary line to the posterior axillary line. PF is mobilized en bloc from the transversalis fascia. Once the working space is fully established (Fig. 1h), additional ports are inserted under laparoscopic guidance. Subsequently, the surgical procedure can be performed.

### Statistical analysis

The following demographic and intraoperative variables were recorded: age, gender, body mass index (BMI), and the time of creating retroperitoneal working space which is defined as the time elapsed from the initial incision to the placement of all trocars. Complications were classified according to the Clavien-Dindo system.

Median and inter-quartile range (IQR) was used for continuous variables. Categorical variables were reported as percentages. Statistical analysis was performed by SPSS software version 22.0.

## Results

Patient demographic and clinical characteristics are detailed in Table 1. Using the laparoscope, we were able to adequately dissect the retroperitoneal space and obtain a satisfactory surgical field for all patients. The median time of creating retroperitoneal working space was 6 (IQR:5,7) min. All procedures were completed successfully. At a median follow-up of 9 (IQR:7,15) months, no dissection-related complications were observed.

**Table 1.**
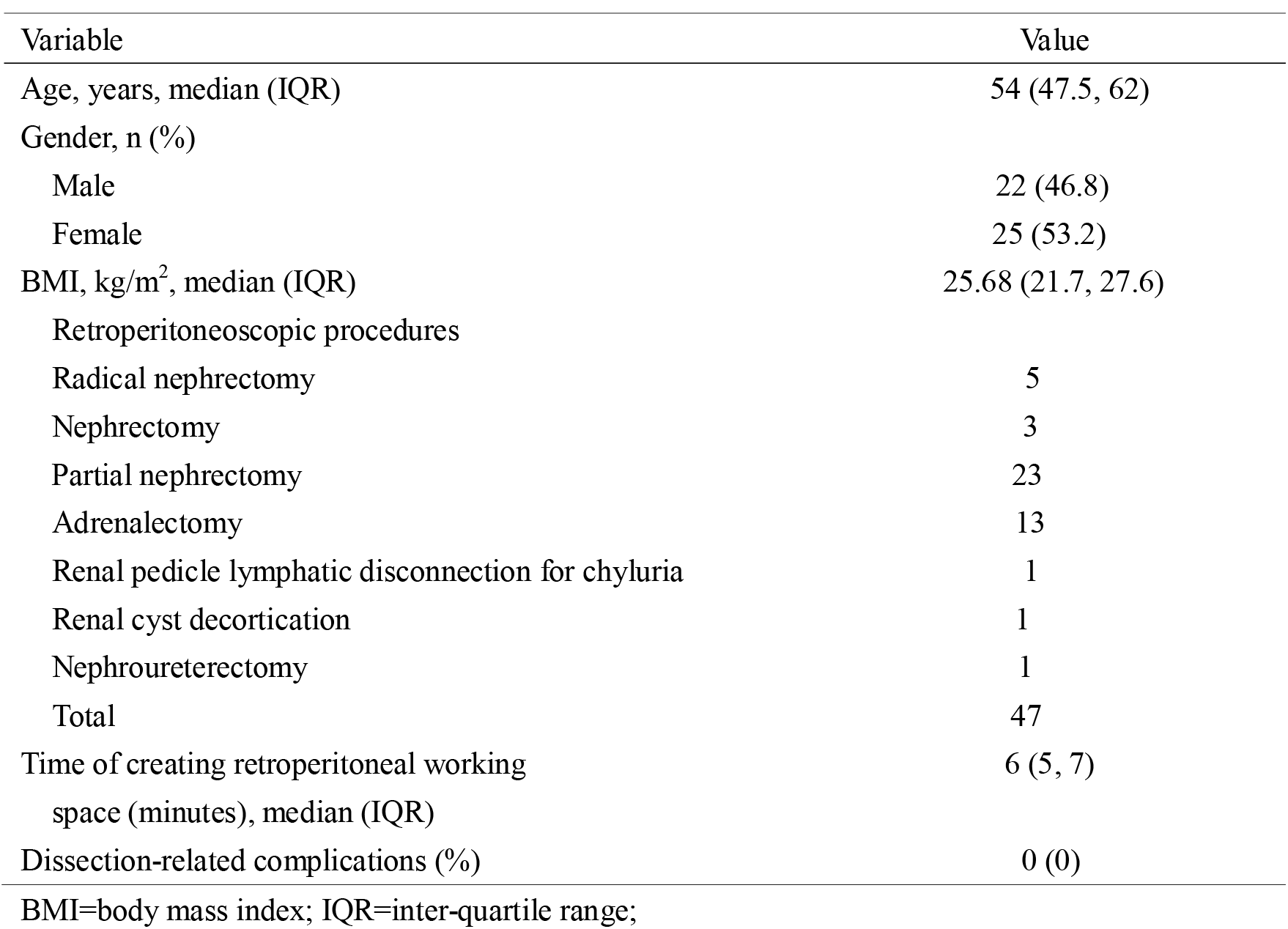
Demographic and clinical characteristics of the 47 patients with modified dissection technique. Variable Value.

## Discussion

Currently, the Hasson and Veress needle techniques are the two major approaches used for retroperitoneal access and creating space for retroperitoneal laparoscopic surgery.

The Hasson technique is commonly used, wherein initial access and balloon placement for retroperitoneal laparoscopy are established using an open approach [1-4]. A 1.5-2.0 cm incision is made just below the tip of the 12^th^ rib in the posterior axillary line, and the lumbodorsal fascia and muscle fibers are bluntly separated using artery forceps [1,3]. Thereafter, the peritoneum is digitally pushed forward to create a retroperitoneal space. Lastly, a balloon is inserted to fully dilate the retroperitoneal space.

Using this technique, intracorporeal manipulation is not performed under direct vision and it is associated with the risk of improper dilation between muscle layers caused by balloon misplacement. Intramuscular dilation can lead to musculature tearing and bleeding, possibly resulting in conversion to an open procedure or postoperative hernia [4]. Furthermore, if the balloon is inserted too deep and dilates within the Gerota fascia, the renal vein can tear [5]. These complications can be avoided by direct visualization using a commercially available transparent balloon where the laparoscope is inserted into the balloon and the proper position of the balloon is confirmed endoscopically. However, this results in higher costs. Another alternative is the use of ultrasound to confirm the position of the balloon [6]. In addition, even if the incision is as small as possible, air leakage around the balloon trocar remains a problem because the index finger is thicker than the 10-mm trocar. Thus, airtight instruments or sutures must be adopted to ensure an airtight incision [2].

During the Veress needle technique, the Veress needle is inserted and followed by “blind” trocar puncture at the midaxillary or posterior axillary lines through a 10-mm skin incision. However, this procedure is associated with several complications [7]. First, the fibroareolar tissue binds to the PF in the retroperitoneum and can not be sufficiently broken down by CO_2_ insufflation via the Veress needle alone. It is thus difficult to obtain an adequate working space, and breaking the fibroareolar septa via laparoscopy is needed. Second, the tip of the trocar will be inserted into the PF (between the psoas muscle posteriorly and Gerota’s fascia anteriorly) or perirenal fat (within Gerota’s fascia) due to the “blind” puncture [8]. Further laparoscopic dissection is performed within fatty tissues, and en bloc fat is separated into several parts, which is often accompanied by vascular breakage. Lastly, laparoscopic visibility is often obscured by blood and fat tissue, and this technique is associated with frequent smudging of the laparoscope and a lack of anatomical landmarks [9]. An insufficient endoscopic view may result in complications, such as pneumothorax [6,7].

Because of the disadvantages of the existing dilation techniques, a modified technique for creating a retroperitoneal working space was introduced at our institution. In this procedure, initial access is obtained by trocar puncture through a 10-mm skin incision without the use of digital dissection. Therefore, air leakage due to a larger incision can be avoided. The tip of the trocar is adjusted to the plane between the TAM and the PF under laparoscopic monitoring. This is important because this layer is composed of semi-transparent loose and flimsy fibroareolar tissue and is nearly avascular. This facilitates a clear laparoscopic view during dissection and en bloc dissection of the PF from the transversalis fasci.

After proper layer entry, dissection movements of the laparoscope are performed. The different visual characteristics of the transversus abdominis muscle, PF, and fibroareolar tissue act as laparoscopic anatomical landmarks, which guarantees that the correct layer is dissected. In our experience, proper layer entry can fill areolar tissue with gas, which contributes to the en bloc mobilization of the PF from the transversalis fasci.

Furthermore, both the tented portion of the flank abdominal wall and the light spot on the skin from the laparoscope could indicate the precise position of the dilation. These factors also contribute to the avoidance of mis- or overdistention. Ultimately, a clear laparoscopic view with rare hemorrhage is possible.

This modified technique offers several unique advantages. First, the skin incision is small, and no blunt dissection of the fascia and muscle with artery forceps is performed. Therefore, intraoperative air leakage is avoided. Mattress sutures or airtight systems are not necessary. Second, a preliminary blunt dissection is performed between the PF and transversalis fasci and not inside the PF. This ensures a clean laparoscopic view because this layer is a relatively avascular semi-transparent reticular fibrous structure. Lastly, manipulation of the entire process is performed using endoscopic monitoring. Therefore, the complications associated with balloon misplacement can be avoided.

## Conclusion

This modified technique is simple and requires limited experience and technical skills. The procedure is performed under distinct endoscopic monitoring, which minimizes the risk of injury from obscured vision or “blind” manipulation. It not only provides a clear working space but also facilitates further dissection of the PF en bloc off Gerota’s fascia. In summary, the modified dilation technique was initially proven safe and effective.

## Data Availability

All data produced in the present study are available upon reasonable request to the authors

## Author contributions

X.F.X. conceived the idea, initial drafting of the article. Cheng: Data collection and data analysis, initial drafting of the article. Z.L., C.J.S., and J.P.D. Data collection. X.M.Z. Interpretation of data, critical revision of the article. J.P.G. Interpretation of data, critical revision of the article.

